# Minimum Dietary Diversity and Associated Factors among Lactating Mothers in Ataye District, North Shoa Zone, Central Ethiopia: A Community-based Cross-sectional Study

**DOI:** 10.1101/2020.10.17.20214320

**Authors:** Lemma Getacher, Gudina Egata, Tadesse Alemayehu, Agegnehu Bante, Abebaw Molla

## Abstract

**Background:** Low dietary diversity superimposed with poor quality monotonous diets is a major problem that often results in undernutrition, mainly micronutrient deficiencies. However, there is limited evidence on minimum dietary diversity and associated factors among lactating mothers in resource-poor settings, including the study area. Therefore, the objective of the study is to assess the prevalence of minimum dietary diversity and associated factors among lactating mothers in Ataye District, Ethiopia.

**Methods:** A community-based cross-sectional study design was used among 652 lactating mothers aged 15-49 years from January 25 to April 30, 2018. Dietary diversity was measured by the minimum dietary diversity indicator for women (MDD-W) using the 24-hour dietary recall method. Data were entered into EpiData version 4.2.0.0 and exported to the statistical package for social science (SPSS) version 24 for analysis using the logistic regression model.

**Results:** The prevalence of minimum dietary diversity among lactating mothers was 48.8%, (95% CI: (44.7%, 52.9%). Having formal education [(AOR=2.16, 95% CL: (1.14, 4.09)], a final say on household purchases [(AOR=5.39, 95% CI: (2.34, 12.42)], home gardening practices [(AOR=2.67, 95% CI: (1.49, 4.81)], a history of illness [(AOR=0.47, 95% CI: (0.26, 0.85), good knowledge of nutrition [(AOR=5.11, 95% CI: (2.68, 9.78)], being from food-secure households [(AOR=2.96, 95% CI: (1.45, 6.07)] and medium [(AOR=5.94, 95% CI: (2.82, 12.87)] and rich wealth indices [(AOR= 3.55, 95% CI: (1.76, 7.13)] were significantly associated with minimum dietary diversity.

**Conclusion:** The prevalence of minimum dietary diversity among lactating mothers was low in the study area. It was significantly associated with mothers having a formal education, final say on the household purchase, home garden, good knowledge of nutrition, history of illness, food-secure households and belonging to medium and rich household wealth indices. Therefore, efforts should be made to improve the mothers decision-making autonomy, nutrition knowledge, household food security and wealth status.

## 1. Background

Appropriate and adequate maternal nutrition is a pivotal public health issue in resource-poor countries throughout the globe, particularly in Africa, Southeast Asia, Latin America, and the Caribbean. Women of reproductive age (WRA) are often nutritionally vulnerable, especially lactating mothers [1]. Lactating mothers, particularly those who live in low-and middle-income countries, are vulnerable to malnutrition due to increased physiological demand, lactogenesis process, eating undiversified monotonous diet and increased nutrient needs during lactation.

Nutritional requirements during lactation are greater than during pregnancy, and they need at least two extra meals per day. When these needs are not met, mothers may suffer from malnutrition especially, from micronutrient deficiencies. To prevent and correct nutritional vulnerability in women of reproductive age, especially during the most 1000 critical days, one of the best nutritional interventions is dietary diversification. Dietary diversity (DD) refers to an increase in the variety of foods across and within food groups capable of ensuring adequate intake of essential nutrients that can promote good health, physical and mental development of women. Moreover, it is a qualitative measure of food consumption that reflects household access to a variety of foods and is also a proxy for the micronutrient adequacy diet of individuals [2-6].

Micronutrient deficiency (aka hidden hunger) is one form of undernutrition that occurs due to insufficient intake, poor knowledge about nutrition, inadequate dietary diversity or sufficient intake combined with impaired absorption due to infection, disease or inflammation and phytates (anti-nutrients) that can decrease the bioavailability of nutrients such as minerals [7]. The eleven micronutrients that are the most important nutrients during lactation are vitamin A, vitamin C, thiamine, riboflavin, niacin, vitamin B6, folate, vitamin B12, calcium, iron, and zinc. Undiversified diet during lactation leads to these important micronutrients being low in breast milk and reflects the poor nutritional status and inadequate dietary intake of micronutrients [2, 8-10].

Low dietary diversity is one of the most important causes of both macronutrient and micronutrient deficiencies [2, 13]. Studies conducted in some Asian countries showed that low dietary diversity ranges from 55.2% to 87.8% [14, 15], whereas in African country studies, low dietary diversity ranges from 42.1% to 88.7% [13, 16, 17]. In Ethiopia, the low dietary diversity ranges from 56.4% to 99.9% [15, 18-20].

The major causes of inadequate dietary diversity stem from wider contextual factors such as lack of education, poor socioeconomic status, cultural beliefs, poor agricultural practices, poverty, disparities in households and food insecurity. Low dietary diversity has both short-term consequences, such as mortality, morbidity, disability and long-term consequences, such as stunting, impaired cognitive ability, poor economic productivity, poor reproductive performance, increased metabolic and cardiovascular diseases, and intergenerational consequences. Hence, the consequences of micronutrient malnutrition not affect only the health and survival of women but also their children [7, 11].

The three main strategies that have been documented for the prevention and control of hidden hunger are short-term supplementation, medium-term food fortification and a long-term focus on dietary diversification [21]. According to different scientific pieces of evidence, there is a growing worldwide consensus to increase the dietary diversity of women (consuming more food groups), especially lactating mothers, which can help to improve their diet quality and infants. Since no single food group can contain all nutrients required for the healthy functioning and performance of the body, there is, therefore, the need for more food groups to be included in the daily diet to meet their nutrient requirements [1, 3, 9]. The government of Ethiopia also has a goal to reduce maternal undernutrition through dietary diversity intervention by launching a revised national nutrition program (NNP II) among women of reproductive age, including lactating mothers and their children in 2016 [22]. Therefore, assessing the prevalence of minimum dietary diversity and associated factors among lactating mothers is vital to improving the quality of diet taken by lactating mothers.

Despite having many studies done elsewhere in the globe on the prevalence of minimum dietary diversity among pregnant women and household level, there is a paucity of data concerning the nature and relative magnitude of minimum dietary diversity among lactating mothers in Ethiopia, particularly there is a lack of study in central parts of the country including the study area. In addition, most of the previous studies performed across the globe, including Ethiopia, did not use the newly recommended indicator, which is called minimum dietary diversity-women (MDD-W), except the South African study, to assess dietary diversity of mothers. Furthermore, previous studies did not address the prevalence of minimum dietary diversity at the community level using this indicator. Therefore, this study was aimed at assessing the prevalence of minimum dietary diversity and associated factors among lactating mothers in Ataye District, North Shoa Zone, Amhara Regional State, Central Ethiopia, including variables that were not included in the previous studies.

## 2. Methods

### 2.1. Study setting, period and design

A community-based cross-sectional study was conducted from January 25 to April 30, 2018 in Ataye District. Ataye is located 270 km away from Addis Ababa, the capital city of Ethiopia. The District has 30 kebeles (lowest administrative unit in Ethiopia). The district has seven health centers, 24 health posts, and one District hospital. The common agricultural practices in the District are *teff*, wheat, sorghum, barley, maize, peas and beans, and some vegetables, such as cabbage, potato, pumpkin, tomato, onion, carrots, and lettuce, and some fruits, such as orange, papaya, banana, and mango, were among others. Lactating mothers aged 15-49 years and lived for at least six months and above in the District during the study period were included in the study, while mothers who were not able to respond to an interview and who had an unusual dietary intake in the previous 24 hours, such as feasts and celebrations, were excluded from the study.

### 2.2. Sample Size Determination and Sampling Procedures

The sample size was estimated using two approaches based on the objectives of the study. To determine the prevalence of minimum dietary diversity, a single population proportions formula was used with the following assumptions: the proportion of maternal dietary diversity among lactating mothers was 43.6% [20], Z_a/2_ had a 95% confidence level of 1.96, a margin of error of 0.05, a non-response of 10%, and a design effect of 1.5. Accordingly, the calculated sample was 624. On the other hand, to assess the predictors of minimum dietary diversity using a double proportion formula considering different predictors significantly associated with the outcome variable, such as educational status, occupational status and meal frequency, were used.

During calculation, assumptions such as a two-sided confidence level of 95%, a margin of error of 5% and power of 80%, a 1:1 proportion of exposed to unexposed ratio, a respective odds ratio for each factor, a non-response of 10% and a design effect of 1.5 were considered using the Epi Info 7 software program, and then the calculated sample size was 647. The latter sample size was considered to increase the power of the study. However, the total number of mothers in the selected kebeles was 652. Thus, due to the nature of the cluster sampling method, all mothers were included in the selected cluster using a cluster sampling technique considering kebeles as clusters.

This study was conducted in two urban kebeles and four kebeles of rural in the District. The numbers of lactating mothers found in each kebele were taken from a family folder documented by the Health Extension Workers (HEWs) with respect to their household. After that, all lactating mothers in randomly selected clusters were included in the study through the house-to-house visit.

### 2.3. Data Collection Methods and Instruments

A structured interviewer-administered questionnaire was first prepared in English and was translated to Amharic (the language spoken in the study area) for data collection and then translated back to English by an independent language expert to ensure its consistency.

Six female grade ten completed students who were fluent speakers in the local language were recruited to participate in data collection. Two-degree health professionals were recruited for the supervision of the data collection procedure. The data collectors underwent a community-based face-to-face interview using a structured and pretested Amharic questionnaire. The mothers were interviewed (on average 30 minutes) in a private setting inside their own house.

### 2.4. Measurements

In this study, the outcome variable (minimum dietary diversity) was measured by minimum dietary diversity for a woman (MDD-W), which is a dichotomous indicator/tool that has been developed by FAO. A total of 16 food groups were considered in this study (i.e. cereals, white tubers and roots, vitamin A-rich vegetables and tubers, dark green leafy vegetables, other vegetables, vitamin A-rich fruits, other fruits, organ meat, flesh meat, eggs, fish and seafood, legumes, seeds and nuts, milk and milk products, oils and fats, sweets, spices, condiments, and beverages). These food groups were further regrouped into ten food groups (i.e. all starchy staples, pulses (beans, peas, and lentils), nuts and seeds, all dairy, flesh foods (including organ meat), eggs, vitamin A-rich dark green leafy vegetables, other vitamin A-rich vegetables and fruits, other vegetables and other fruits) during analysis [3, 4].

The Minimum Dietary Diversity Score (MDDS) was calculated for each lactating mother during the previous 24 hours to classify mother’s dietary diversity as adequate (≥5 food groups) or not adequate (<5 food groups) from ten food groups. Then, the outcome variable was coded as minimum dietary diversity score ≥5 food groups, as “1” and minimum dietary diversity score <5 food groups as “0” for logistic regression analysis.

Household food insecurity was measured with the Household Food Insecurity Access Scale (HFIAS), a structured, standardized and validated tool that was developed mainly by FANTA, to classify households as food secure or not [24, 25]. A scale is to be a valid tool in measuring household food insecurity among both rural and urban areas of Ethiopia with Cronbach’s alpha values of 0.76 for round 1 and 0.73 for round 2 [26]. In this study, nine standard Household Food Insecurity Access Scale (HFIAS) questions adapted from FANTA were computed. All “Yes” responses were coded “1” and “No” responses were coded “0”, and the responses were summed to obtain the household food insecurity status. The HFI status, which had a high internal consistency (Cronbach’s alpha = 0.927), was further dichotomized as “food secure” and “food insecure households which coded as “1” and as “0”, respectively, during analysis. Furthermore, the household wealth index was analyzed using principal component analysis (PCA) by using approximately 20 locally available household assets considering the assumptions.

The knowledge of mothers about nutrition was computed based on six questions that included awareness of the mothers about nutrition and dietary diversity practice. The mothers who scored above the mean cut-off point were considered to have good knowledge and coded as “1”, whereas those who scored below this cut–off point were considered to have poor knowledge and coded as “0”.

### 2.5. Data Quality Control

The adapted and further developed English language questionnaire was translated and contextualized into the Amharic language by an Amharic language speaker who has attended a Master of Arts in the Amharic language. It was translated back to the English language by a person who attended Master of Arts in the English language, and comparison was made on the consistency of the two versions. Data were collected using the translated and pretested Amharic version structured questionnaire. The questionnaire was modified further after a pretest was conducted. During data collection, the interviewers informed mothers about all the details of the research. The mothers were encouraged to feel free and told that the confidentiality of their responses was assured and that no information was shared with anybody without the investigator. After this, women who were willing to participate and signed the informed consent document were interviewed.

Two days of training (both theoretical and practical) was given for six female grade ten completed students and the two supervisors. The training was focused on interview technique, ethical issues, rights of the participants, reading through all the questions and understanding them and ways of decreasing under/over-reporting and maintaining confidentiality.

Three weeks before the actual data collection, the questionnaire was pretested outside to the selected kebele on 5% of the total sample size to ensure the validity of the tool. After the pretest was performed, all the necessary adjustments were made. To minimize bias, interviews were conducted in an area with adequate confidentiality, privacy and without the involvement of any other person other than the respondent. On-site supervision was carried out during the whole period of data collected daily by the supervisors and principal investigators. At the end of each day, questionnaires were reviewed and cross-checked for completeness, accuracy, and consistency by the supervisor and principal investigator, and in the end, corrective measures were taken.

### 2.6. Statistical Analysis

All the interviewed questionnaires were checked visually by the principal investigator. Data were coded, entered and cleaned using EpiData version 4.2.0.0 software. Double data entry was performed by two data clerks to cross-check the data for completeness before analysis. The entered data were exported and analyzed with Statistical Package for Social Science (SPSS) version 24 software. Simple descriptive statistics, such as simple frequency distribution, measures of central tendency, measures of variability and percentages, were performed to describe the demographic, socioeconomic, and maternal-related characteristics of the respondents. Then, the information was presented using tables and figures. The continuous variables were tested for normal distribution using a histogram, Q-Q plot and some statistical tests such as Kolmogorov-Smirnov and Shapiro-Wilk tests.

Bivariable analysis and crude odds ratio along with a 95% confidence interval (CI) were used to see the association between each independent variable and the outcome variable by using binary logistic regression. Independent variables with a p-value of ≤ 0.25 were included in the multivariable analysis to control for confounding factors. Multicollinearity was checked to see the linear correlation among the independent variables by using standard error (SE). Variables with a standard error of ≥ 2 were dropped from the multivariable analysis. The fitness of the model was tested by Hosmer-Lemeshow’s goodness-of-fit test model coefficient with an enter method, which was found to be insignificant with a large P-value (P=0.374) and the Omnibus tests were significant (P=0.0001). The adjusted odds ratio along with 95% CI was estimated to identify the factors associated with dietary diversity using multivariable logistic regression analysis. All tests were two-sided, and the level of statistical significance was declared at a p-value less than 0.05.

The ethical approval letter was obtained from Haramaya University, College of Health and Medical Science Institutional Health Research Ethics Review Committee (IHRERC). The approval letter was dated 15 January 2018 and numbered Ref C/AC/R/D/897/18. Before informed consent was obtained, a clear description of the study title, purpose, procedure, duration, possible risks and benefits of the study was explained for each study participant. Their rights during the interview were guaranteed. Then written and signed informed consent was obtained from each respondent before starting the interview. Questionnaire code numbers were used to maintain the confidentiality of information gathered from each study participant throughout the study.

## 3. Results

### 3.1. Sociodemographic Characteristics of Study Participants

Out of 652 planned lactating mothers, 631 of them aged 15-49 years participated in a response rate of 96.8%. The mean age (± SD) of lactating mothers was 30.48 (±6.36) years. From the total of participating, respondents, 417 (66.1%) were rural by residence, 345 (54.7%) were in the age group 15-30, 545 (86.4%) were Orthodox Christian by religion, 566 (89.7%) were Amhara by ethnicity and 501 (79.1%) were married by their marital status (Table 1).

**Table 1:**
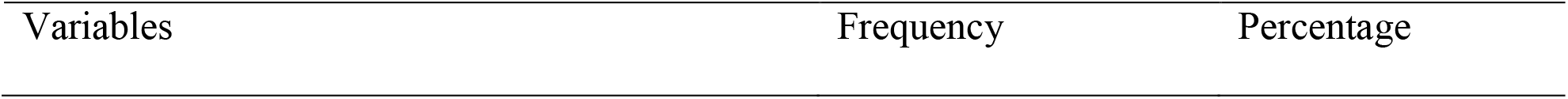
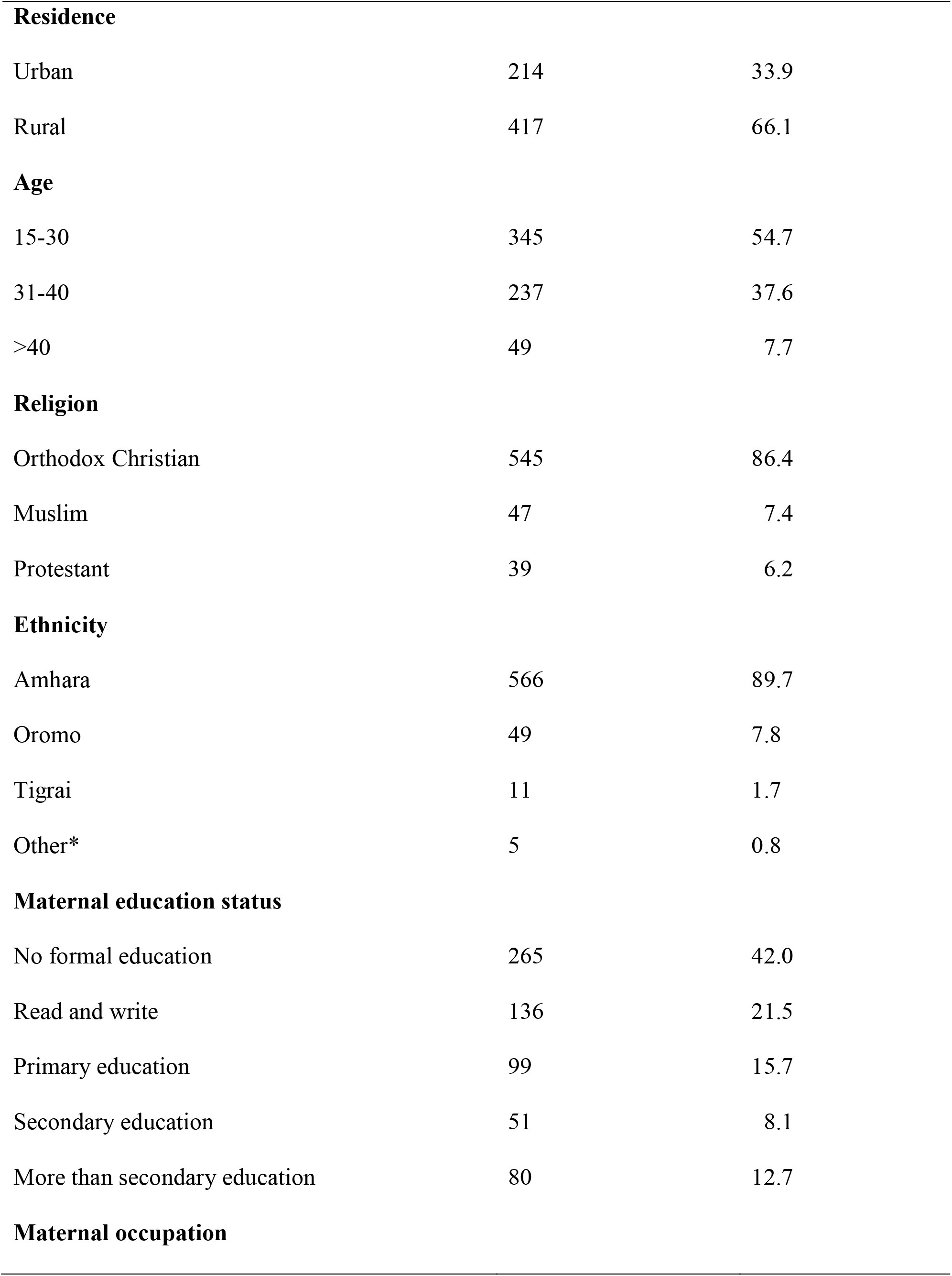
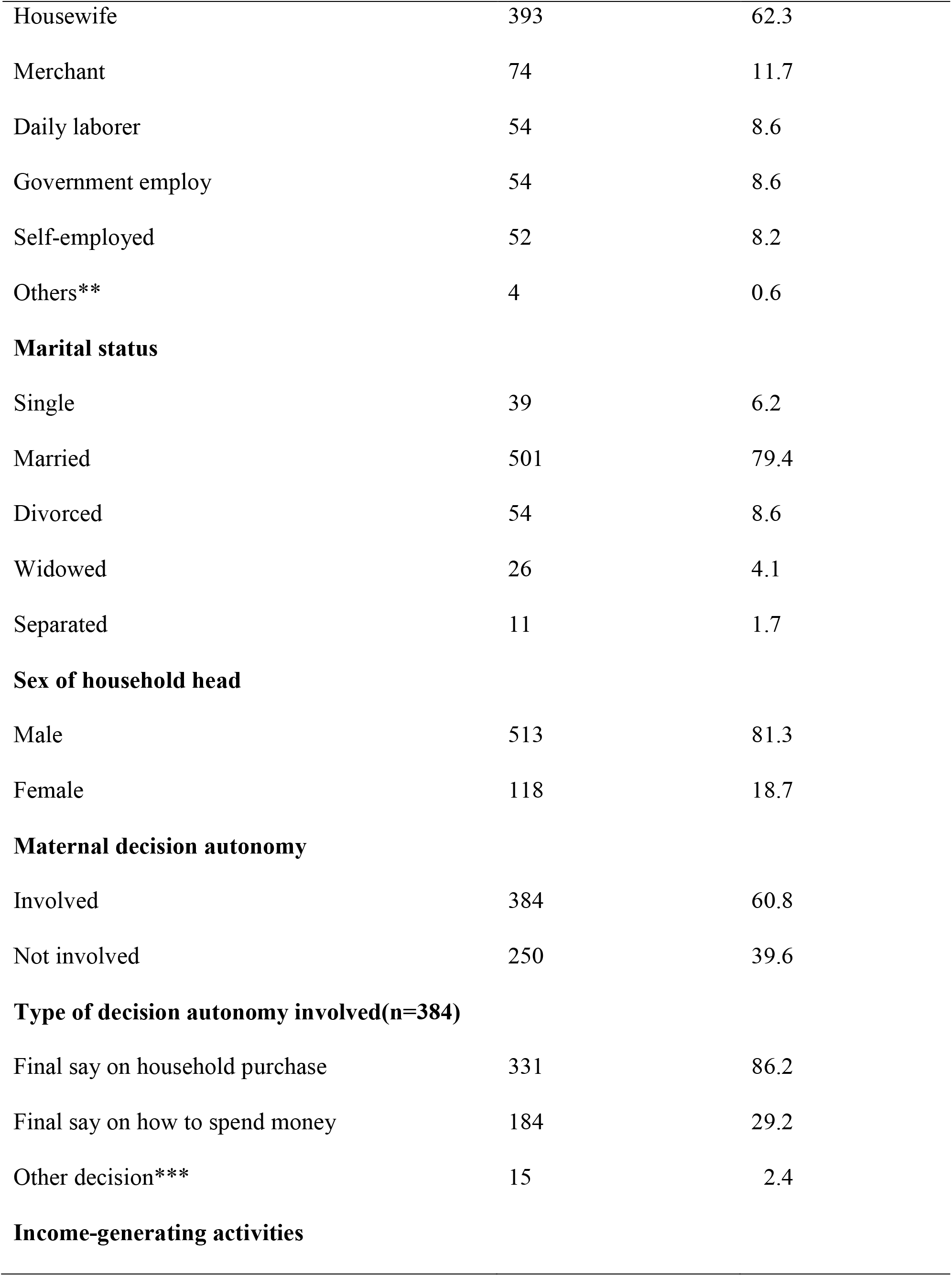
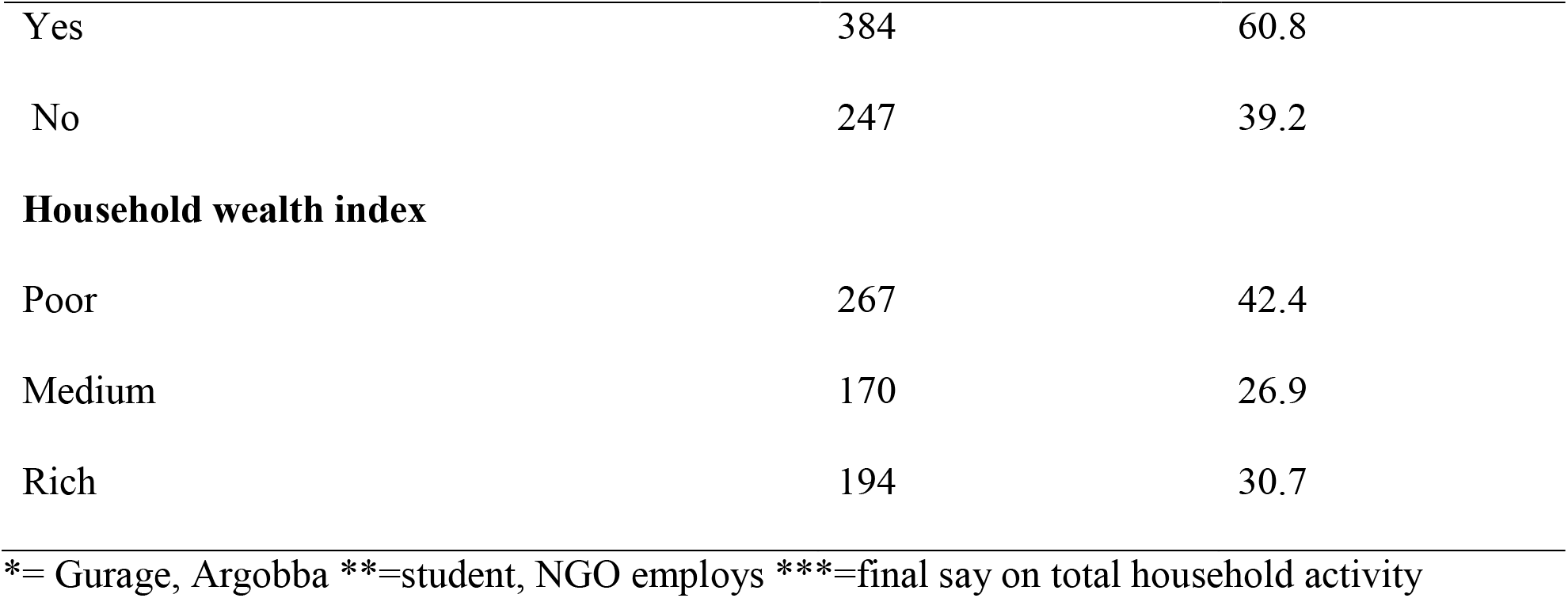
Sociodemographic characteristics of lactating mothers in Ataye District, Central Ethiopia, 2018 (n = 631)

| Variables | Frequency | Percentage |
| --- | --- | --- |
| **Residence** |  |  |
| Urban | 214 | 33.9 |
| Rural | 417 | 66.1 |
| **Age** |  |  |
| 15-30 | 345 | 54.7 |
| 31-40 | 237 | 37.6 |
| >40 | 49 | 7.7 |
| **Religion** |  |  |
| Orthodox Christian | 545 | 86.4 |
| Muslim | 47 | 7.4 |
| Protestant | 39 | 6.2 |
| **Ethnicity** |  |  |
| Amhara | 566 | 89.7 |
| Oromo | 49 | 7.8 |
| Tigrai | 11 | 1.7 |
| Other* | 5 | 0.8 |
| **Maternal education status** |  |  |
| No formal education | 265 | 42.0 |
| Read and write | 136 | 21.5 |
| Primary education | 99 | 15.7 |
| Secondary education | 51 | 8.1 |
| More than secondary education | 80 | 12.7 |
| **Maternal occupation** |  |  |
| Housewife | 393 | 62.3 |
| Merchant | 74 | 11.7 |
| Daily laborer | 54 | 8.6 |
| Government employ | 54 | 8.6 |
| Self-employed | 52 | 8.2 |
| Others** | 4 | 0.6 |
| **Marital status** |  |  |
| Single | 39 | 6.2 |
| Married | 501 | 79.4 |
| Divorced | 54 | 8.6 |
| Widowed | 26 | 4.1 |
| Separated | 11 | 1.7 |
| **Sex of household head** |  |  |
| Male | 513 | 81.3 |
| Female | 118 | 18.7 |
| **Maternal decision autonomy** |  |  |
| Involved | 384 | 60.8 |
| Not involved | 250 | 39.6 |
| **Type of decision autonomy involved(n=384)** |  |  |
| Final say on household purchase | 331 | 86.2 |
| Final say on how to spend money | 184 | 29.2 |
| Other decision*** | 15 | 2.4 |
| **Income-generating activities** |  |  |
| Yes | 384 | 60.8 |
| No | 247 | 39.2 |
| **Household wealth index** |  |  |
| Poor | 267 | 42.4 |
| Medium | 170 | 26.9 |
| Rich | 194 | 30.7 |

\*= Gurage, Argobba \*\*=student, NGO employs \*\*\*=final say on total household activity

### 3.2. Maternal Health-care and Feeding Practice

More than half of lactating mothers 354 (56.1%) had one to three frequency of ANC visits in their previous/last pregnancy. Among those who had ANC visits, 297 (47%) delivered in the health center and 119 (18.9%) delivered in a hospital (Table 2).

**Table 2:** Maternal healthcare and feeding practices of lactating mothers in Ataye district, Central Ethiopia, 2018 (n = 631)

| Variables | Minimum Dietary Diversity |  | Frequency (%) |
| --- | --- | --- | --- |
|  | ≥5 FGs (%) | <5 FGs (%) |  |
| **Number of ANC visit** |  |  |  |
| No visit | 32 (5.1) | 87 (13.8) | 119 (18.9) |
| 1-3 | 174 (27.6) | 180 (28.5) | 354 (56.1) |
| ≥4 | 102 (16.2) | 56 (8.8) | 158 (25.0) |
| **Place of delivery** |  |  |  |
| Home | 78 (12.4) | 137 (21.7) | 215 (34.1) |
| Health center | 144 (22.8) | 153 (24.2) | 297 (47.0) |
| Hospital | 86 (13.6) | 33 (5.3) | 119 (18.9) |
| **Postnatal follow-up** |  |  |  |
| Yes | 189 (57.3) | 141 (42.7) | 330 (52.3) |
| No | 119 (39.5) | 182 (60.5) | 301 (47.7) |
| **Source of food** |  |  |  |
| Own production | 146 (23.1) | 219 (34.7) | 365 (57.8) |
| Purchasing | 157 (24.9) | 99 (15.7) | 256 (40.6) |
| Food aid/relief | 5 (0.8) | 3 (0.5) | 8 (1.3) |
| Other sources* | 1 (0.15) | 1 (0.15) | 2 (0.3) |
| **Home gardening practice** |  |  |  |
| Yes | 179 (28.4) | 81 (12.8) | 260 (41.2) |
| No | 129 (20.4) | 242 (38.4) | 371 (58.8) |
| **Meal frequency** |  |  |  |
| Two meals only or below | 21 (3.3) | 32 (5.1) | 53 (8.4) |
| Three meals | 197 (31.2) | 237 (37.6) | 434 (68.8) |
| Three meals above | 90 (62.5) | 54 (37.5) | 144 (22.8) |
| **Illness in the last 2 weeks** |  |  |  |
| Yes | 74 (11.7) | 169 (26.8) | 243 (38.5) |
| No | 234 (37.1) | 154 (24.4) | 388 (61.5) |

\*=no formal source like helping from others/relatives, FGs= food groups

### 3.3. Minimum Dietary Diversity Score of lactating mothers

The prevalence of the minimum dietary diversity score (≥5 food groups) among lactating mothers was 48.8 (95% CI: (44.7%, 52.9%) during the preceding 24 hours of the survey. The majority of the study participants had consumed all starchy staple foods (99.4%) and pulses (beans and peas) (79.1%). The least consumed food group was other fruits (8.7%) (**Figure 1**).

**Figure 1:**
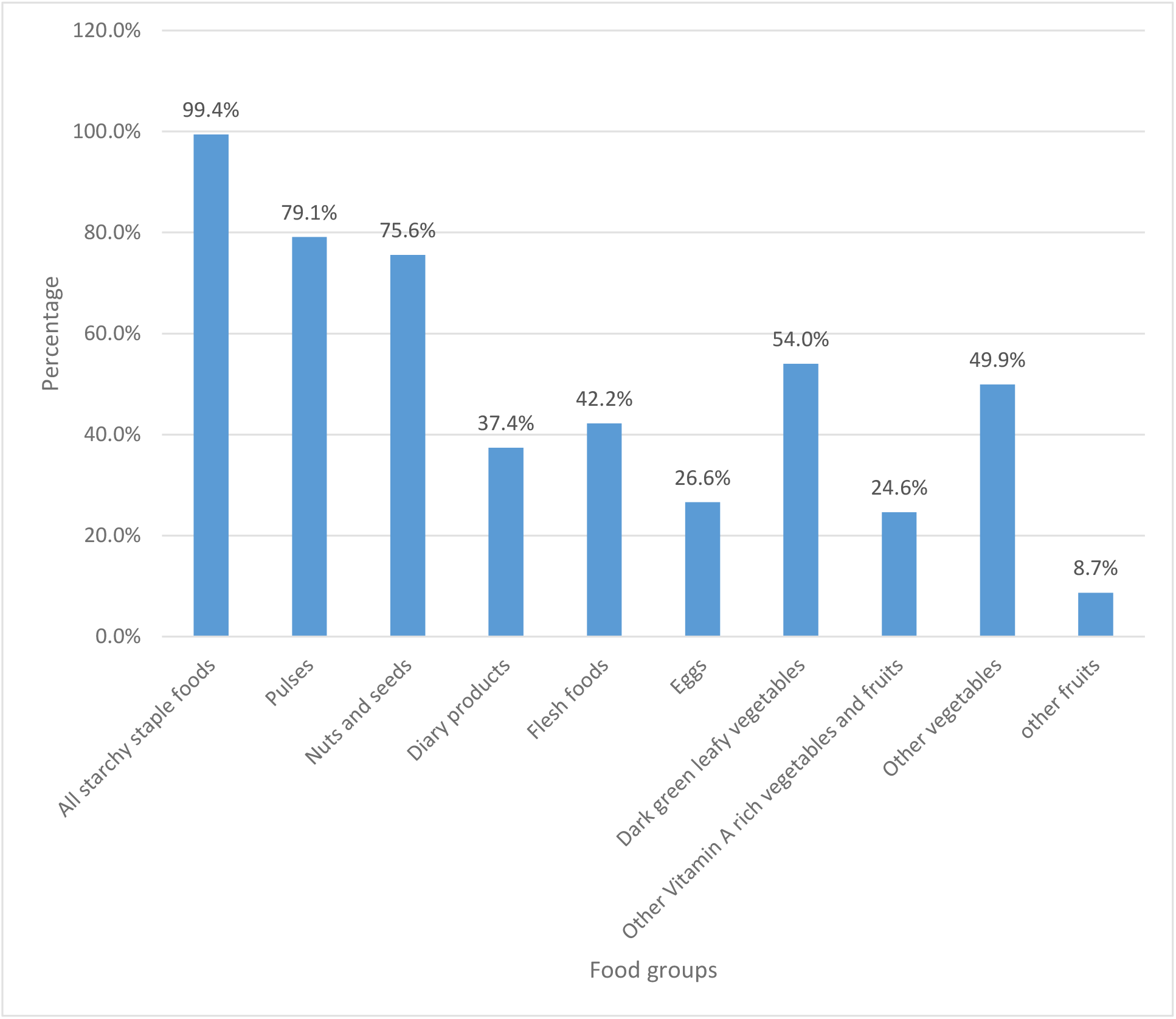
The ten grouped food groups consumed by lactating mothers in Ataye District, Central Ethiopia, 2018 (n=631)

### 3.4. Knowledge about nutrition

Regarding knowledge about nutrition, of the total participants in this study, 54.8% (95% CI: 50.9, 58.5) had good knowledge of nutrition.

### 3.5. Household food security status

The household food security status of the study participants was 75.9%, 95% CI: (72.4, 79.3) food secure, while 24.1%, 95% CI: (20.7, 27.6) were food insecure households.

### 3.6. Factors Associated with Minimum Dietary Diversity among Lactating Mothers

In the bivariable logistic regression analysis model, educational status of lactating mothers, mothers who had the final say on a household purchase, mothers who had income-generating activities, increased number of ANC visits, place of delivery, postnatal follow-up, the main source of food, home gardening practice, history of illness, daily eating pattern, knowledge of nutrition, household food insecurity, and household wealth indices were significantly associated with minimum dietary diversity of mothers and became candidates for further multivariable analysis.

In the multivariable logistic regression analysis model, those variables with a P-value of ≤ 0.25 in the bivariable analysis were entered into a multivariable analysis using the enter method to identify the independent predictors of the minimum dietary diversity score. After adjusting for all possible confounders, the formal educational status of mothers, maternal decision-making autonomy related to a final say on a household purchase, home gardening practice, good knowledge of nutrition, illness in the last two weeks, food-secure households and medium and rich household wealth indices remained significant and independent predictors of the minimum dietary diversity score of mothers.

Lactating mothers who had formal education were more than two times [(AOR=2.16, 95% CL: (1.14, 4.09)] more likely to have minimum dietary diversity scores than those who had no formal education. Regarding maternal decision-making autonomy, mothers who had the final say on household purchases were more than five times [(AOR=5.39, 95% CI: (2.34, 12.42)] more likely to achieve minimum dietary diversity than their counterparts. The odds of minimum dietary diversity scores were nearly three times higher among mothers who had home gardening practice [(AOR=2.67, 95% CI: (1.49, 4.81)] than their counterparts.

Mothers who had a history of illness in the last two weeks were 53% [(AOR=0.47, 95% CI: (0.26, 0.85) less likely to reach a minimum dietary diversity score than their counterparts. The odds of minimum dietary diversity score were more than five times [(AOR=5.11, 95% CI: (2.68, 9.78)] more common among mothers who had good knowledge of nutrition than those who had poor knowledge of nutrition. Food-secure households had nearly three times the minimum dietary diversity scores than food-insecure households [(AOR=2.96, 95% CI: (1.45, 6.07)].

Mothers from households who had medium and rich wealth indices were nearly six times and more than three times more likely to have minimum dietary diversity experience than poor households [(AOR=5.94, 95% CI: (2.82, 12.87)] and [(AOR= 3.55, 95% CI: (1.76, 7.13)], respectively (Table 3).

**Table 3:**
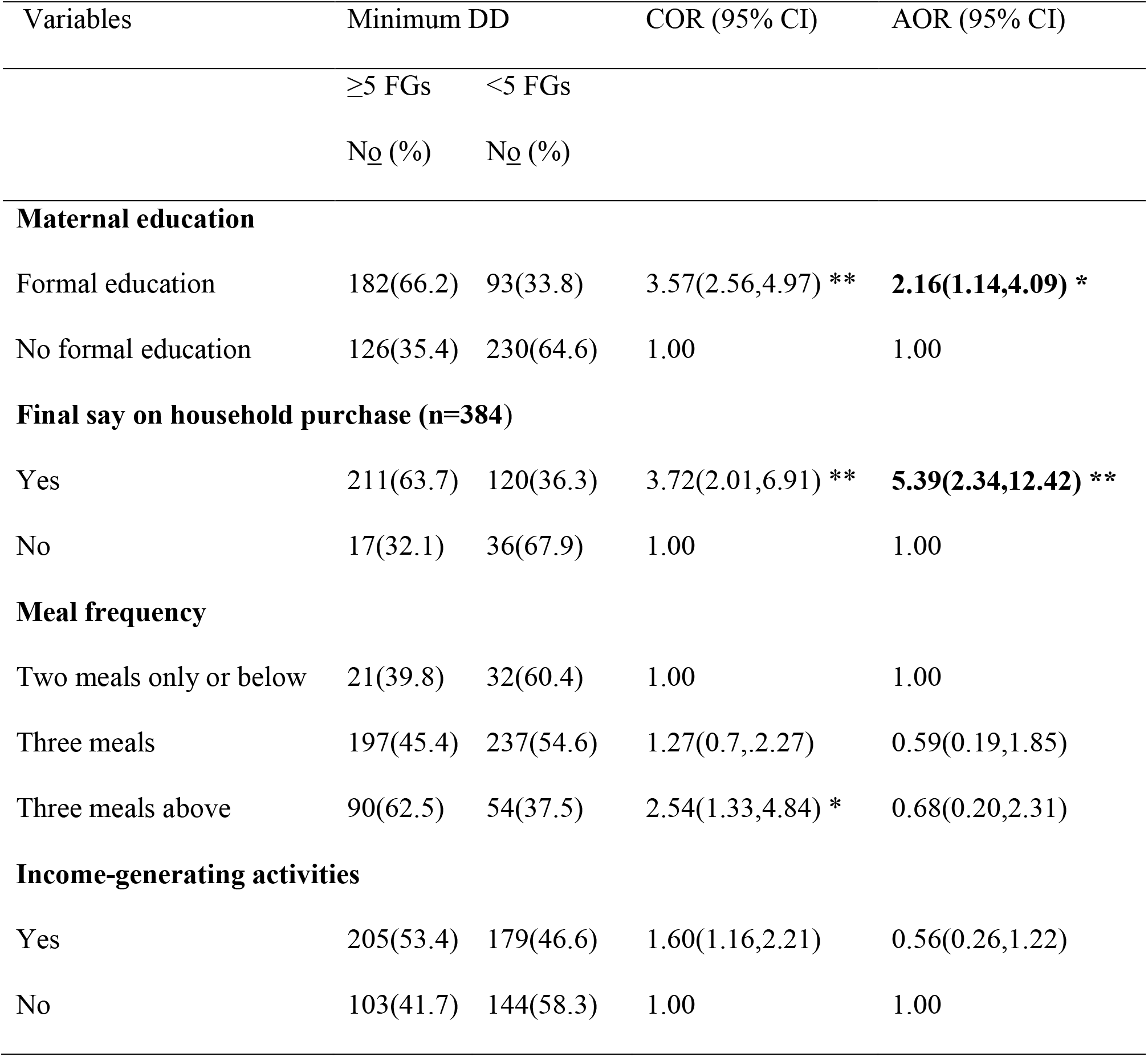
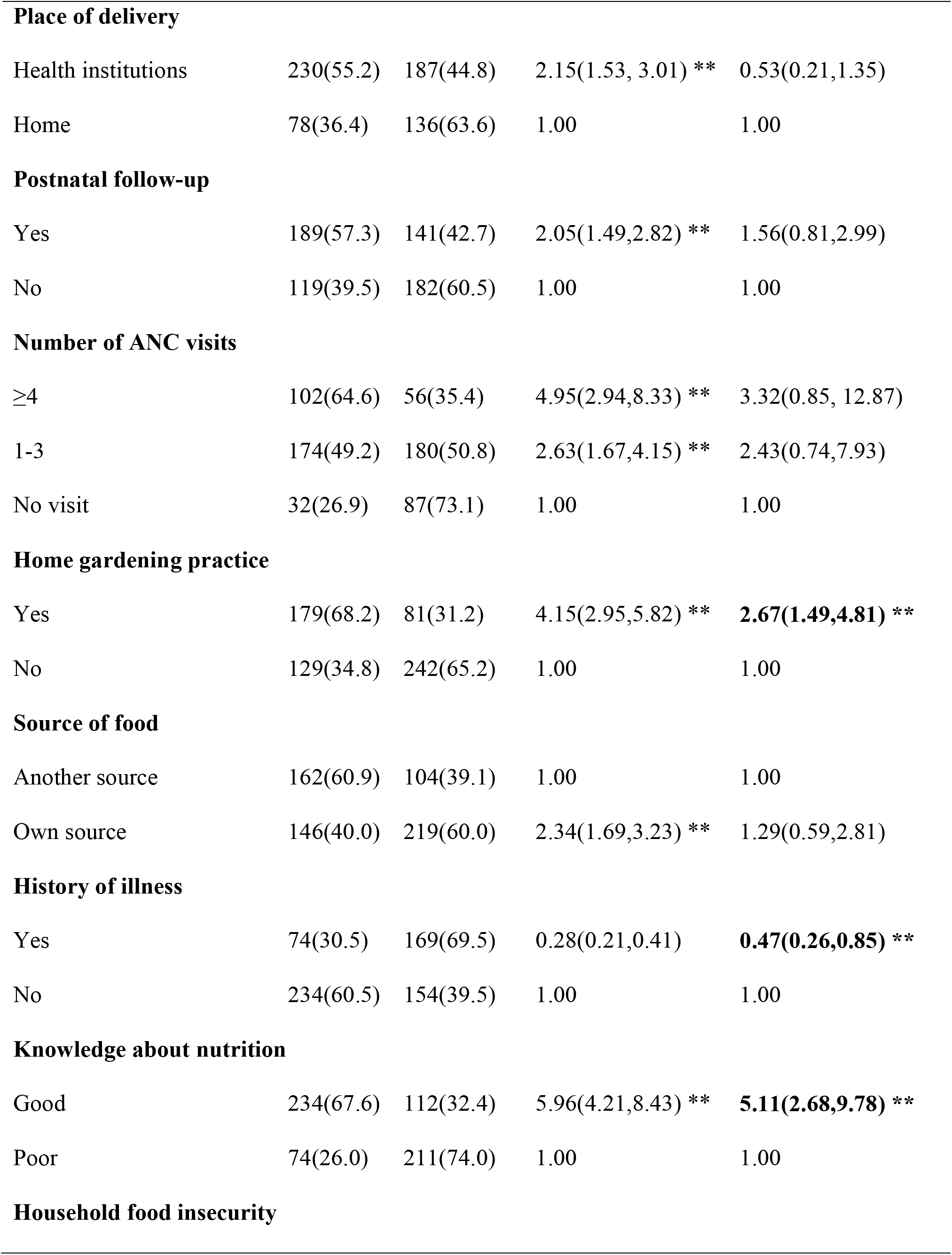
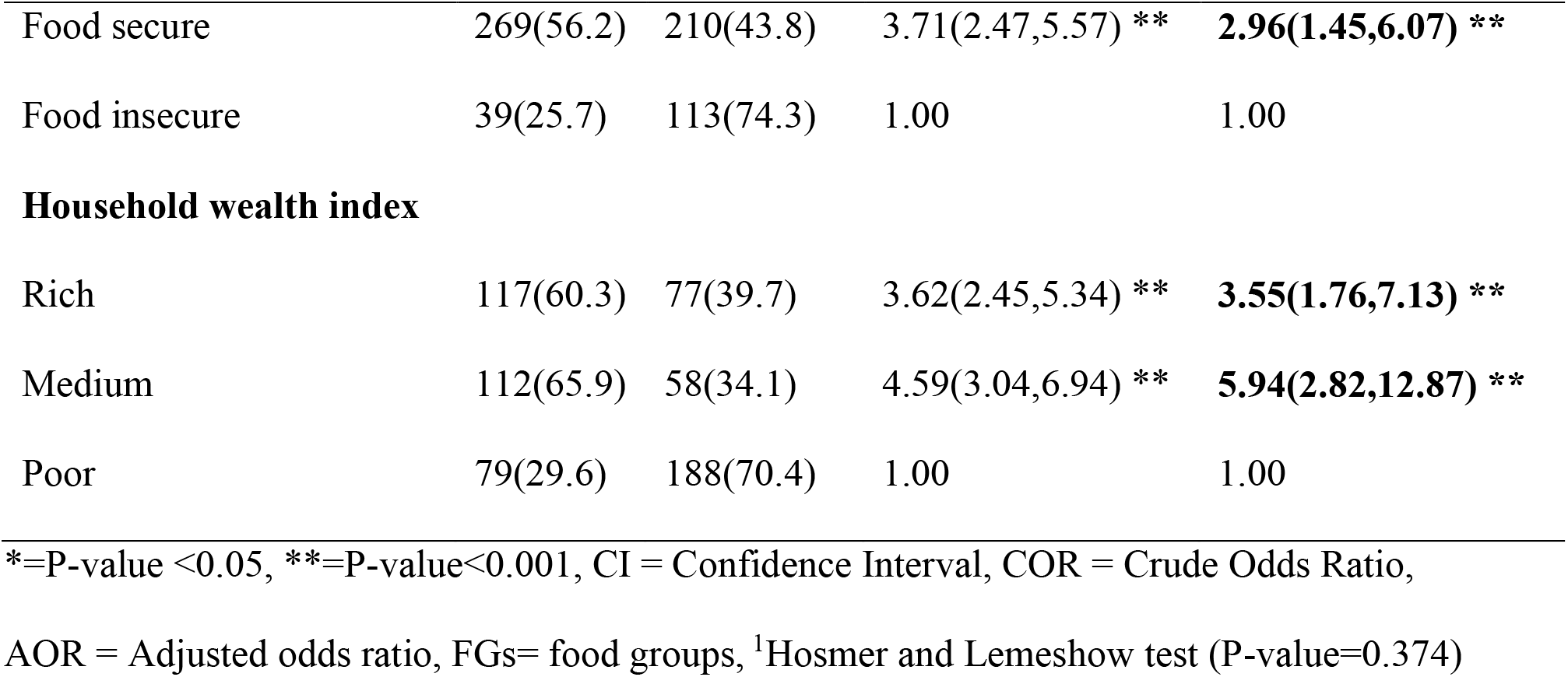
Bivariable and multivariable logistic regression1 analysis of factors associated with MDD among lactating mothers in Ataye district, Central Ethiopia, 2018 (n=631).

## 4. Discussion

In this study, approximately 48.8% of lactating mothers achieved a minimum dietary diversity score, and they were more likely to have higher (more adequate) micronutrient intakes than 51.2% of lactating mothers who did not. This result indicated that the prevalence of minimum dietary diversity in the study area is low. The 2016 FAO guideline recommended that all of the mothers should achieve the minimum dietary diversity score (≥5 food groups) to have adequate micronutrient adequacy (3). The present study also examined the demographic and socioeconomic characteristics, household-related characteristics, maternal healthcare and feeding practice-related characteristics associated with minimum dietary diversity among lactating mothers in Ataye District. Accordingly, attending formal education, having decision-making autonomy on household purchases, having home gardening practices, having good knowledge of nutrition, having an illness in the last two weeks, being from food secure households and being in medium and rich household wealth indices were independent predictors of minimum dietary diversity.

The prevalence of minimum dietary diversity is relatively higher than the study findings carried out elsewhere in the world such as Bangladesh (12.8%) and Vietnam (22%) [15], South Africa (25%) [16], Ghana (43.1%) [13], Aksum Town (43.6%) [20] and Jimma (32.8%) [18]. In contrast, this prevalence is lower than that in the study performed in Uganda (57.9%) [17]. This inconsistency could be due to the difference in the sample size because some of the studies were large surveys, they had a relatively large sample size (study done in Bangladesh and Vietnam) and some of the studies also had a lower sample size than the present study (study done in South Africa and Aksum). The study period and seasonal variation (since this study was done during the postharvest season) may also be another explanation for the observed discrepancy in which it can result in food security status change, while most of the previous studies had a high prevalence of food insecurity compared to the present study, which is nearly one-fourth. Methodological (measurement tool) differences may be another possible explanation because the current study uses the newly recommended indicator of ten food groups rather than the nine food groups’ indicators. The feeding culture, educational status and socioeconomic status of the study participants were also other explanations for this observed variation.

The educational status of mothers was associated with a minimum dietary diversity score of lactating mothers. Lactating mothers who had formal education were more than two times highly dietary diversified mothers than those who had no formal education. This finding is similar to that of prior studies done in Ghana [13] and three countries (Bangladesh, Vietnam, and Ethiopia) [15]. This could be related to the fact that educated mothers may obtain sufficient information about varieties of food during their health service utilization and understanding the importance of increasing dietary diversity and meal frequency during lactation than non-educated mothers in case they would have the probability of taking a variety of food groups.

Minimum dietary diversity was associated with maternal decision-making autonomy practice. Mothers who had the final say on household purchase were more than five times more likely to achieve minimum DDS compared to those who had no final say on household purchases. This figure is in line with the finding of Ghana, women who had a final say were almost twice as likely to achieve a higher DD score [13]. This consistency may be because as the mother’s involvement is increased in the family on a nutrition-related decision, they have the probability of purchasing and deciding on an important variety of foods for the households, their child and themselves, as a result, they may have increased diversify diets.

In the present study, the home gardening practice of lactating mothers was found to be an independent predictor of the minimum DDS of mothers. Lactating mothers who practice home gardening showed nearly three-fold higher likelihood of consuming highly diversified diets than their counterparts. This is comparable with the study conducted in Aksum town, Ethiopia [20], which demonstrated that home gardening was strongly associated with diet diversity. This might be because mothers from households with gardens would grow vegetables and fruits and become beneficiary from gardens to diversify their daily food intake; therefore, they may obtain additional diet options that enhance the diversity of the household’s food consumption of different food groups in their diets.

Good maternal knowledge of nutrition predicted the minimum dietary diversity score of lactating mothers in the current study area. Mothers who scored above the mean cut-off point upon responding to nutrition and food diversity knowledge questions were more than five times more likely to achieve a minimum dietary diversity score compared to their counterparts. This result is comparable with the study done in three countries (Bangladesh, Vietnam, and Ethiopia) and Uganda [5, 17]. This may be because mothers who had increased knowledge about nutrition may know the importance of varieties of foods and have a greater likelihood of taking these varieties of food groups. It could also be related to mothers getting enough nutrition and dietary diversity-related information and education when they have good access to health services during their antenatal and postnatal follow up. This helps them to increase their knowledge about varieties of food groups, and as a result, they may have adequate intake and consumption.

Maternal history of illness was associated with minimum DDS of mothers. Mothers who had a history of illness were 53% less likely to have the probabilities of achieving minimum DD scores when compared to their counterparts. A possible explanation could be that illness may decrease appetite and intake of the diet which in turn affects the dietary intake and dietary diversity of mothers. In addition, most of the respondents (nearly two-thirds) in the present study were rural residents and they may not receive enough health service in the nearby health facility immediately as they worsned, leading to a decrease in the minimum DDS mothers.

Another important predictor of the minimum dietary diversity score of mothers was household food insecurity. Lactating mothers from food-secure households had nearly three-fold higher probabilities of achieving a high level of MDDS than food-insecure households. This is supported by previous studies in three countries Bangladesh, Vietnam, and Ethiopia [15]. This could be explained in such a way that food security promotes the intake of adequate quantity and quality of diet that in turn contributes to having a better minimum dietary diversity score and nutritional status.

The odds of MDD score was more common among mothers found in medium and rich wealth indices. Mothers belonging to medium and rich socioeconomic status households were nearly six times and four times more likely to have minimum dietary diversity scores compared to poor socioeconomic status households respectively. This finding is consistent with previous results in three countries (Bangladesh, Vietnam, and Ethiopia) [15], South Africa [16] and Ghana [13]. The possible explanation behind this may be that as the socioeconomic status of the family increases, they would have stable food security status in case their probabilities of eating diversified food groups would also increase.

In summary, this finding will also support one of the strategic objectives of the National Nutrition Program II (NNP-II) in Ethiopia, which was launched to improve the nutritional status of women of reproductive age, including adolescent girls, pregnant women, and lactating mothers, through appropriate dietary diversity to alleviate the current burden of micronutrient deficiencies [22].

Regarding the strength and limitations of the study, the major strength of the current study is that using the newly recommended 10-point food group indicator, MDD-W, instead of the previous 9-point food group using a community-based cross-sectional study design was the limitation of other studies. However, recall bias was one of the limitations of the study since some of the questions we asked about the event that occurred 24 hours and 4 weeks back. This was minimized by probing the respondents about the event. Seasonal variation was also another limitation of the study. This study was also affected by reporting and social desirability bias. Therefore, this study could not reject the possibility of reporting bias from lactating mothers who did not eat. To minimize these biases lactating mothers were asked in a private setting.

## 5. Conclusion

Nearly half of the lactating mothers achieved the minimum dietary diversity score in the current study. Regarding the predictors of minimum dietary diversity score, attending formal education, having decision autonomy on the household purchase, having home gardening practice, having good knowledge of nutrition, having illness in the last two weeks, being food secure household and being in medium and rich household wealth indices were the significant and independent predictors of minimum dietary diversity. Therefore, the government of Ethiopia, particularly the Ataye District Health Office, should be prioritizing, planning, designing and initiating dietary diversity intervention programs aimed at improving maternal nutrition through appropriate food-based approaches. Strengthen nutrition education programs on proper maternal MDD practices and adequate dietary intake during lactation to improve the nutrition outcomes of lactating mothers. Cooperatively work with health extension workers to increase the awareness of lactating mothers on how to improve the dietary diversity practice of mothers. Create strong multisectoral collaboration targeted at improving the mother’s educational status, decision-making autonomy (women empowerment) and family socioeconomic status. District Health Extension Workers (HEWs) should also facilitate and promote nutrition education about dietary diversification for lactating mothers and the community at large. Educate mothers about visiting health service institutions when they become sick and about increasing their intake of varieties of food. Work in cooperation with developmental agents to increase the minimum dietary diversity score of lactating mothers in the District. Local NGOs should actively work in collaborating with the woreda health office to increase the awareness and knowledge of lactating mothers about food diversity and nutrition. Collaboratively work with another stakeholder to increase the stability of the HFS and socioeconomic status of the mother’s family and the whole community to prevent inadequate DD practice of the mother’s, thereby preventing poor nutritional status. Finally, the scientific community should study a different study design, such as a prospective cohort study design, that is recommended to address seasonal variability during the preharvest and postharvest seasons. Since dietary diversity is multifactorial and multifaceted, further study is needed to identify other independent predictors for lactating mothers’ dietary diversity.

## Data Availability

The data used to support the findings of this study are available from the corresponding author upon reasonable request.

## Abbreviations

DDS: Dietary Diversity Score
EDHS: Ethiopian Demographic and Health Survey
FAO: Food and Agricultural Organization of the United Nation
FANTA: Food and Nutrition Technical Assistant
HFIAS: Household Food Insecurity Access Scale
MDDS: Minimum Dietary Diversity Score
MDD-W: Minimum Dietary Diversity for Women
WHO: World Health Organization

## Author’s contribution

LG: - participated in the design of the study, performed the data collection, performed the statistical analysis and served as the lead author of the manuscript. GE: - participated in the design of the study, revised subsequent drafts of the paper and contributed to the finalization of the manuscript. TA, AB, and AM: - participated in the design of the study, statistical analysis and finalization of the manuscript. All authors read and approved the final manuscript.

## Acknowledgments

We would like to express our deepest gratitude to Haramaya University, College of Health and Medical Science for ethical clearance. Our great thanks also deserve to our study participants, data collectors, supervisors and language translators for their invaluable contribution to this study.

